# Connectome-based predictive modeling of brain pathology and cognition in Autosomal Dominant Alzheimer’s Disease

**DOI:** 10.1101/2024.09.01.24312913

**Authors:** Vaibhav Tripathi, Joshua Fox-Fuller, Vincent Malotaux, Ana Baena, Nikole Bonillas Felix, Sergio Alvarez, David Aguillon, Francisco Lopera, David C Somers, Yakeel T. Quiroz

## Abstract

**INTRODUCTION:** Autosomal Dominant Alzheimer’s Disease (ADAD) through genetic mutations can result in near complete expression of the disease. Tracking AD pathology development in an ADAD cohort of Presenilin-1 (*PSEN1)* E280A carriers’ mutation has allowed us to observe incipient tau tangles accumulation as early as 6 years prior to symptom onset.

**METHODS:** Resting-state functional Magnetic Resonance Imaging (fMRI) and Positron-Emission Tomography (PET) scans were acquired in a group of *PSEN1* carriers (n=32) and non-carrier family members (n=35). We applied Connectome-based Predictive Modeling (CPM) to examine the relationship between the participant’s functional connectome and their respective tau/amyloid-β levels and cognitive scores (word list recall).

**RESULTS:** CPM models strongly predicted tau concentrations and cognitive scores within the carrier group. The connectivity patterns between the temporal cortex, default mode network, and other memory networks were the most informative of tau burden.

**DISCUSSION:** These results indicate that resting-state fMRI methods can complement PET methods in early detection and monitoring of disease progression in ADAD.

## 1. Introduction

A rare form of AD is genetically determined through autosomal dominant mutations in the *APP*, *PSEN1*, or *PSEN2* genes. One of the most well-characterized autosomal dominant AD (ADAD) mutations is Presenilin1 (*PSEN1*) E280A, which causes early-onset AD dementia with nearly 100% certainty [1,2]. Studies on this ADAD cohort established that *PSEN1* E280A carriers develop Mild Cognitive Impairment (MCI) and dementia around the ages of 44 and 49 respectively [3], with the buildup of cerebrospinal fluid (CSF) amyloid-β starting around 19 years prior to the onset of the disease [1]. Whereas late-onset AD is associated with reduced clearance of amyloid-β, ADAD is more related to the overproduction of Aβ_42_ [2]. Studies have also shown the relationship between amyloid-β and tau deposition in ADAD subjects [4]. Longitudinal studies have shown that the neocortical Aβ_42_ accumulation starts around 16 years before onset and precedes tau accumulations in the entorhinal cortex. Tauopathy then reaches the neocortex and is followed by hippocampal atrophy and subsequent cognitive decline [5]. Elevated levels of tau deposition were observed within medial temporal lobe regions in amyloid-β positive *PSEN1* carriers 6 years before clinical onset of AD [4].

Advances in PET imaging have allowed the estimation of amyloid-β concentrations associated with AD pathology in the brain [6]. In recent years, tau pathology has been increasingly associated with brain hypometabolism, patterns of neurodegeneration, and clinical manifestation and is related to the changes due to amyloid-β accumulation [4,7]. These advances have resulted in predicting subsequent progression towards dementia. PET scans are the only imaging modality allowing the localization of AD pathology *in-vivo*. However, it is expensive, not readily available, and involves the injection of a radioactive substance which has minimal risk for normal subjects but elevated risks for a few patient populations like with types of lymphoma [8]. Functional Magnetic Resonance Imaging (fMRI) is more readily available and can be possibly used to complement PET-based measures to estimate the effect of increased AD pathology on brain function [9].

Functional connectivity computed through resting-state fMRI has emerged as a robust method to track brain integrity [10] and parcellate the brain into various brain networks [11–13]. Cerebral blood flow is altered in AD, especially in the temporal-parietal-occipital cortex [14]. This hypometabolism is associated with changes in functional connectivity that can be tracked using resting-state scans. Studies showed that amyloid-β and tau pathologies are associated with brain network segregation and integration in ADAD subjects [9] and changes in functional connectivity between the posterior cingulate cortex and medial temporal lobe [2]. Though individuals share common patterns of functional brain organization, they also have idiosyncratic differences. Many fMRI studies average data across the subject pool to improve the signal-to-noise ratio, but this results in the loss of information about individual variations [15–18]. There have been recent efforts to further consider individual differences in fMRI studies [19]. One such method, called Connectome-based Predictive Modeling (CPM) [20], allows researchers to make predictions of a range of cognitive measures like fluid intelligence and attention. [21,22]. Although brain-wide association studies may require large number of subjects [23], carefully designed protocols can overcome them [24]. Individual AD progressions and neuropathologies are heterogeneous [25–27], hampering early diagnosis and preventative measures [28]. This highlights the need to develop quantitative assays that incorporate the complexity of individual differences.

Relating tau and amyloid concentrations to rsfMRI connectivity can help us develop less expensive and more easily accessible tools for the early detection and diagnosis of AD. In this study, we analyzed the network connectivity of carriers of *PSEN1* E280A mutation from the Colombia-Boston (COLBOS) cohort (n=32) out of which seven had developed MCI, along with age-matched controls (n=35). We first computed the edge connectivity across large-scale brain networks defined using the Yeo 2011 atlas [13]. Second, the edge connectivity was associated with both cortical tau and amyloid-β concentrations and behavioral scores across the two groups using CPM technique. Model performance was tested within and across groups.

## 2. Methods

### 2.1 Participants

The Colombian kindred of about 5000 individuals has a predisposition towards the *PSEN1* gene. About 1500 are carriers and will develop AD with a near 100% certainty. The Massachusetts General Hospital COLBOS longitudinal biomarker study used in this project consisted of 32 *PSEN1 E280A* carriers (including seven MCI) and 35 age-matched family members (details in table 1). The study was approved by the Institutional Ethics Review Board of the University of Antioquia in Colombia and Massachusetts General Hospital in Boston. All subjects gave their informed consent for participation in the study. We used the Spanish versions of the neuropsychological task battery from the Consortium to Establish a Registry for Alzheimer’s Disease (CERAD) including word list learning, delayed recall tasks along with the Mini Mental State Examination (MMSE) [29] as previous studies have shown them to be a robust measure of cognitive decline in this population [30].

**Table 1:** Demographics and cognitive data table.

|  | Mean (SD) |  |  |  |
| --- | --- | --- | --- | --- |
| Parameter | Controls | ADAD Carriers |  |  |
|  |  | All | MCI | Non-MCI |
| N (# Males) | 35 (17) | 32 (13) | 7 (3) | 25 (10) |
| Age | 36.41 (5.64) | 38.31 (6.02) | 45.00 (2.94) | 36.44 (5.29) |
| Education | 10.80 (4.15) | 9.31 (4.16) | 6.57 (4.69) | 10.08 (3.64) |
| MMSE | 28.94 (0.89) | 27.00 (3.10) | 22.00 (2.98) | 28.40 (0.94) |
| WLL | 20.91 (3.10) | 16.88 (5.03) | 10.14 (2.23) | 18.76 (3.84) |
| WL Recall | 7.69 (1.24) | 5.28 (2.99) | 0.71 (0.70) | 6.56 (1.96) |

MMSE – Mini Mental State Examination; WLL – CERAD Word List Learning; WL Recall – Word list recall.

### 2.2 MRI Acquisition

The MRI image acquisition was performed on a Siemens 3T Tim Trio system (Siemens Healthcare, Erlangen, Germany) at Massachusetts General Hospital using a 12-channel head coil. High-resolution T1-weighted MPRAGE sequences were used for anatomical data (voxel size = 1 mm isotropic, 160 slices, TR=2300 ms, TE = 2.98 ms, field of view = 256 mm). Functional BOLD data was collected using T2* gradient-echo echo-planar imaging (EPI) sequences with a whole head coverage (TR = 3 sec, TE = 30 msec, Flip angle = 85°, voxel size = 3 mm isotropic, 47 slices). In the resting-state scan, the subjects were asked to remain still, keep eyes open with normal blinking, and fixate on the cross. Two resting-state runs were collected where each run was six minutes long with 120 TRs with an initial 4 TRs for T1-stabilization.

### 2.3 MRI Preprocessing

MRI datasets were preprocessed using the fMRIPrep software version 22.0.2 [31] which is implemented using Nipype 1.8.5 [32] and utilizes Nilearn across multiple steps [33]. The anatomical data preprocessing included intensity correction, brain tissue segmentation using FSL [34], and the Freesurfer recon-all pipeline to align to surface space. Functional data were skull stripped, motion corrected, slice time corrected, and high-pass filtered (128-sec cutoff). Physiological regressors were extracted to remove noise from the BOLD signals followed by co-registration to the reference space using Freesurfer [35,36] and CIFTI space using Ciftify pipeline [37]. We demeaned the signal across time after the preprocessing pipeline, regressed out the global signal, and concatenated across the runs before further network analysis.

### 2.4 PET Acquisition and preprocessing

The subjects underwent PET imaging at Massachusetts General Hospital. PET data were acquired on a Siemens/CTI ECAT HR Scanner (3D mode; 63 image planes; 15.2 cm axial field of view; 5.6 mm trans axial resolution; 2.4 mm slice interval). [F18] Flortaucipir (FTP) images were acquired after a 9.0 to 11.0 mCi bolus injection between 80 and 100 minutes in 4 separate 5-minute frames. Each subject’s PET data was co-registered to the T1 MPRAGE using SPM8 (https://www.fil.ion.ucl.ac.uk/spm/software/spm8/). The [F18] Flortaucipir (FTP) values were averaged across the Freesurfer ROIs within the Desikan-Killiany atlas (https://surfer.nmr.mgh.harvard.edu/fswiki/CorticalParcellation Freesurfer [35,36] as standardized uptake value ratio (SUVR) followed by 8 mm smoothing using a gaussian kernel. 11C Pittsburgh compound B (PiB) PET images were acquired after an 8.5 to 15 mCi bolus injection with 60-minute dynamic acquisition in 69 frames (12×15 seconds, 57×60 seconds). The data were co-registered to T1 MPRAGE and values were spatially averaged over the Freesurfer ROIs and were expressed as distribution volume ratio (DVR) with cerebellum as the reference ROI.

### 2.5 Connectome-based Predictive Modeling

Connectome-based Predictive Modeling (CPM) is a machine-learning approach to data analysis that seeks to associate patterns of functional connectivity in the brains of individuals with behavioral and/or cognitive measures [20,22]. Here, we adapt this approach to associate MRI functional connectivity with localized PET measurements of tau and amyloid-β deposits in addition to behavioral scores. While large subject pools are typically required to build accurate CPM models linking functional connectivity to cognitive measures, linking two brain measures, namely MRI functional connectivity and PET data appears to have much lower subject pool requirements [38,39]. In our approach, we first selected a measure (e.g., amyloid-β DVR in a particular ROI) that we want to model, one value per subject. We then created a functional connectivity correlation matrix using Spearman correlation coefficients computed with SciPy [40]. Here, we used Schaefer 400 parcellation [12] for the cortex seeds, Yeo-Buckner 17-network atlas for the cerebellum [41] and Tian S1 level atlas [42] for the remaining subcortex which includes 32 nodes, for a total of 449 ROIs. We averaged the resting-state time course data across all vertices or voxels that comprise each seed. We then performed a Pearson correlation of each pair of time courses to create a 449 x 449 functional connectivity matrix for all participants. This matrix represents the individual’s ‘functional connectome’. We then correlated the cognitive measure with each edge correlation value in the functional connectivity matrix across subjects in our training datasets and created a mask for edges displaying significant association using uncorrected p-values < 0.2. We masked the subject’s connectivity matrix and averaged out the positive and negative edges separately, giving one negative and one positive summed correlation value per subject. These correlation values were associated with the behavioral measure and found the relationship between the two using Ordinary Least Squares (OLS) regression as implemented in SciPy. We divided the subjects into training and testing sets, such that a given model was constructed using data from one set of subjects and then applied to a different set of subjects. We used a k-fold approach to make predictions for all subjects. We then used the averaged mask from the k-fold analysis and the averaged OLS model and made predictions on the testing dataset. Therefore, the reported results for a particular prediction reflect the average results across multiple models, where the training and testing subjects have been varied in the k-fold approach. The results stayed consistent with changing the number of folds from 8 to 16 across datasets.

Studies have shown that the progression of the amyloid-β and tau pathologies have separate trajectories with neocortical amyloid-β developing earlier followed by increases in tau in the entorhinal cortex, hippocampus and associated networks [5]. We computed averaged tau and amyloid-β information across the ROIs from the Desikan-Killiany atlas [36]. We then selected different ROIs to model tau and amyloid-β concentrations. The choice of ROIs was defined based on the proteinopathy-specific relevance of these regions to inform on AD progression, as estimated from earlier studies [2]. We ran our CPM model for tau concentrations in the entorhinal cortex, precuneus, hippocampus and inferior temporal (IT) cortex, and separately for amyloid-β concentrations in the Frontal Lateral Retro splenial (FLR) cortex. Separate CPM models were constructed and analyzed for each PET ROI. We also constructed a CPM model linking functional connectome with behavioral measures including CERAD word list learning, delayed recall scores and the MMSE score. We analyzed the accuracy of our models using Pearson correlations between actual and predicted values.

## 3. Results

We applied the CPM approach to form associations between functional connectivity and tau/amyloid/behavioral values across the different groups. Prior research has estimated that amyloid-β accumulates across the neocortex years before symptom onset whereas tau accumulations are observed initially in the entorhinal and trans entorhinal cortex subsequently spreading to various brain regions including the hippocampus, amygdala, and IT [5]. We focused on amyloid-β in the Frontal Lateral Retro splenial (FLR) neocortical areas and tau values in the entorhinal cortex, amygdala, hippocampus, and associated networks. The CPM approach yields two separate networks for each model, one based on the positive connections and the other based on the negative connections. We therefore obtained positive network and negative network predictions for each ROI of interest.

For the *PSEN1* carrier group, we obtained significant predictions for tau concentrations across entorhinal cortex (Figure 1, Positive network: r(30) = 0.53, p = 0.002; Negative network: r(30) = 0.53, p = 0.002), precuneus (Figure 2, Positive network: r(30) = 0.50, p = 0.003; Negative network: r(30) = 0.52, p = 0.002), IT (Figure 3, Positive network: r(30) = 0.51, p<0.003; Negative network: r(30) = 0.57, p < 0.001) and hippocampus (Figure 4, Positive network: r(30) = 0.34, p = 0.054; Negative network: r(30) = 0.38, p = 0.033). The predictions for amyloid-β in FLR cortex had moderate effect size although did not cross statistical significance (Figure 5, Positive network: r(30) = 0.20, p=0.284; Negative network: r(30) = 0.26, p=0.145). We also found significant model accuracy for word list recall cognitive score prediction (Figure 6, Positive network: r(30) = 0.45, p<0.006; Negative network: r(30) = 0.45, p<0.006) as well as MMSE scores (Positive network: r(30) = 0.47, p<0.006; Negative network: r(30) = 0.37, p<0.05) and word list learning scores (Positive network: r(30) = 0.48, p<0.005; Negative network: r(30) = 0.34, p<0.01). In *PSEN1* non-carriers, we found that models were predictive of hippocampal tau (Figure 7, r(33) = 0.36, p<0.033). However, CPM models failed to predict the other regional markers (precuneus tau, entorhinal tau, amyloid FLR) (all p < 0.05). *PSEN1* carriers with MCI were crucial for model success, as models trained across carriers without the MCI subjects had a moderate effect size (r = 0.25-0.4) but failed to reach significance (p > 0.05). Most tau CPM models were successful even without the predictions based on carriers with MCI, but with overall lower prediction accuracies (Entorhinal tau - Positive network: r(23) = 0.45, p = 0.024; Negative network: r(23) = 0.52, p = 0.007; Precuneus tau - Positive network: r(30) = 0.58, p < 0.001; Negative network: r(30) = 0.63, p < 0.0001; IT tau - Positive network: r(23) = 0.27, p = 0.198; Negative network: r(23) = 0.46, p < 0.022), and again the model accuracies failed to reach significance for amyloid-β, word list recall score, and hippocampal tau predictions (r = 0.23-0.33, p > 0.05).

**Figure 1:**
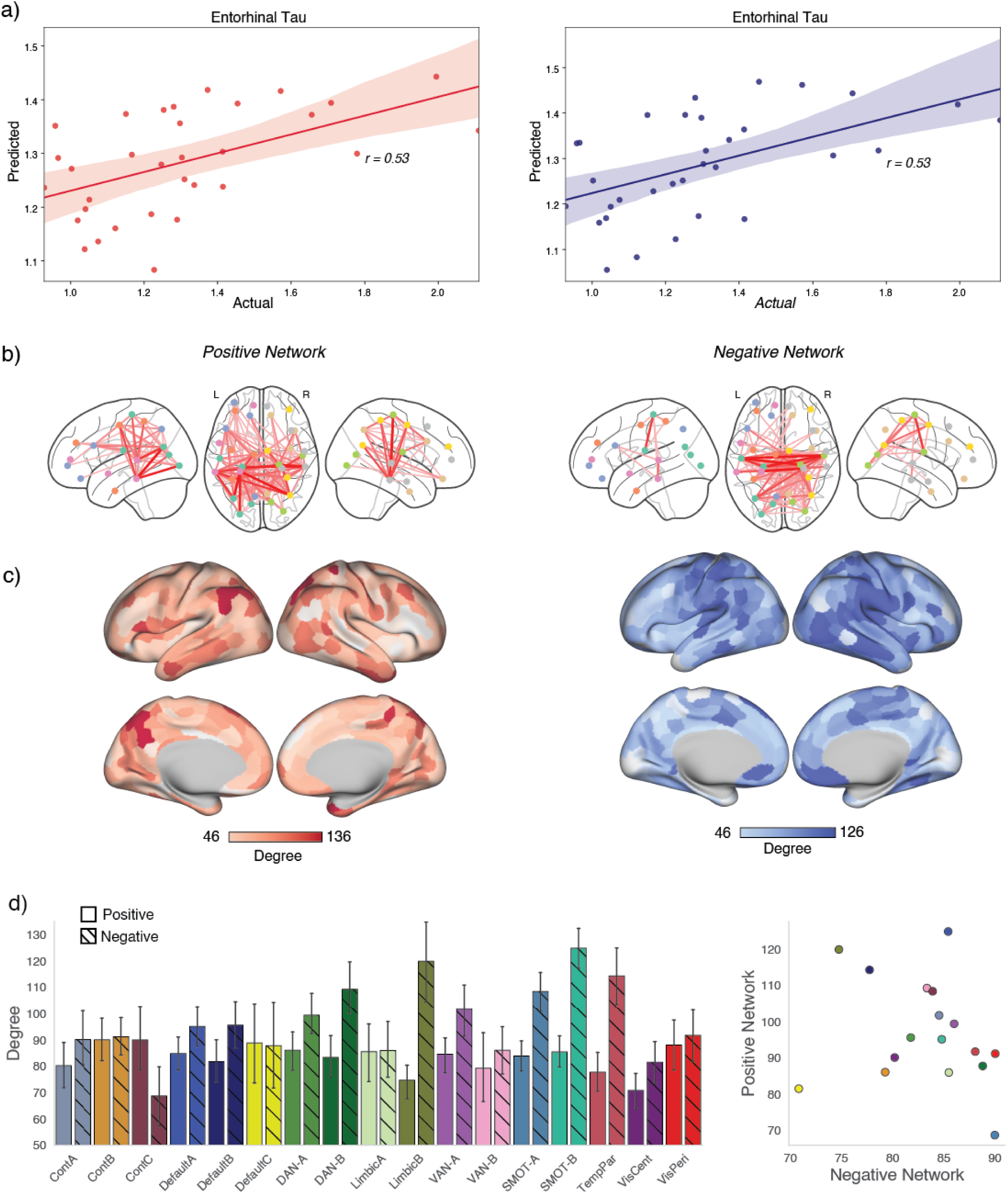
Connectome-based Predictive Modeling for entorhinal tau: a) We trained and predicted the CPM model between functional connectivity and the tau concentrations in the entorhinal cortex within the carriers in the COLBOS dataset. The model was trained using either positive edges (left) or negative edges (right) and equally predicted tau values. b) For the positive network-based model, the connectivity between IPL, posterior cingulate cortex, IT, dorsal attention and temporal cortex is the most predictive of the entorhinal tau values. The bottom panels show the degree of each node in the network model (total of all functional connections, maximum 448) overlayed on the c) cerebral cortex and d) collated across networks represented as a bar plot and scatter plot across the positive and negative networks. Here, the strongest predictors in the model were the DMN, CCN, DAN along with the IPL and PCC regions. For the negative network-based model, the inter-hemispheric connectivity between the somatomotor regions was the most predictive of the tau values. The somatomotor, DAN, temporal regions and parts of the DMN along medial PFC were the most connected regions in the CPM model. The scatter plot shows how different networks (highlighted in Yeo 17 network color scheme) are represented along the positive and negative network models.

**Figure 2:**
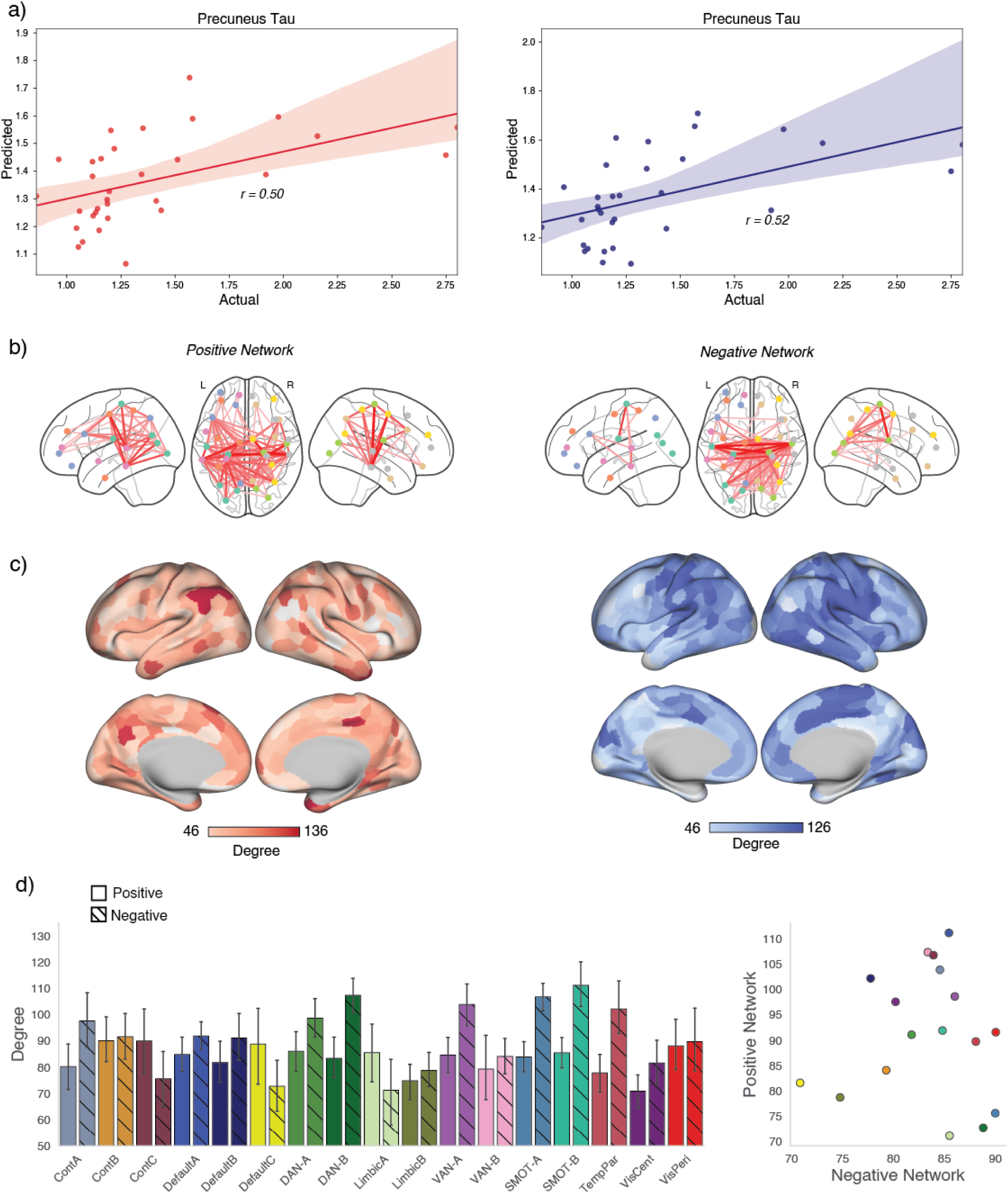
Connectome-based Predictive Modeling for precuneus tau: a) We trained and predicted the CPM model between functional connectivity and the tau concentrations in the precuneus within the carriers in the COLBOS dataset. b) Similar to the entorhinal cortex, the positive network-based model had the connectivity among IPL, PCC, DAN, and IT as the most predictive. c,d) The degree strength of DMN, cognitive control and DAN ROIs along the IPL, PCC regions as the strongest predictors in the model. For the negative network-based model, we found the inter-hemispheric connectivity among the somatomotor, DAN, precuneus, and IT sulcus as strongest predictors.

**Figure 3:**
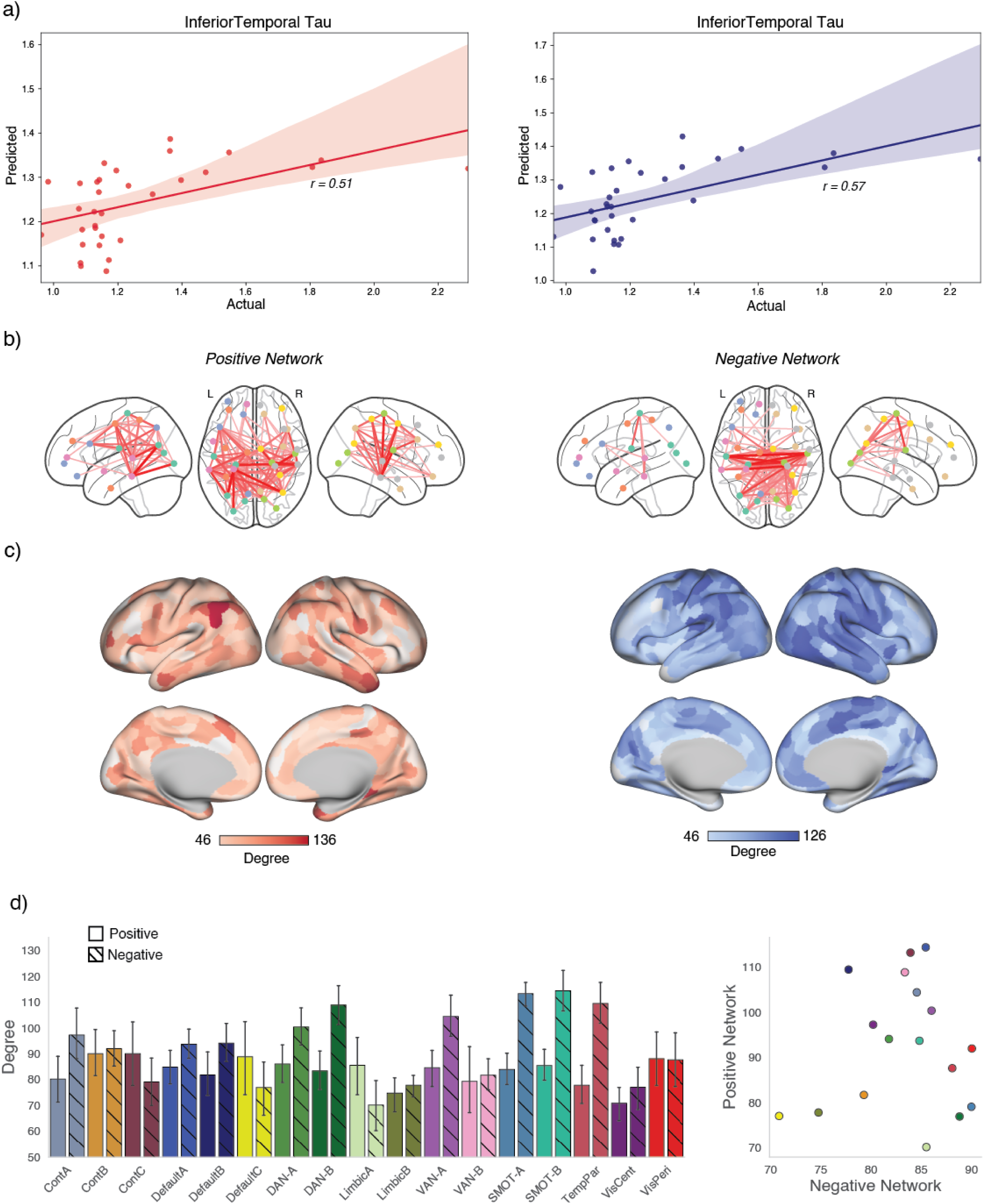
Connectome-based Predictive Modeling for IT tau: a) We trained and predicted the CPM model between functional connectivity and the tau concentrations in the IT within the carriers in the COLBOS dataset. b) For the positive network-based model, the connectivity between IPL, PCC, IT was the most predictive of the IT tau values. For the negative network-based model, the inter-hemispheric connectivity between the somatomotor regions was the most predictive of the tau values. c,d) The somatomotor, DAN, temporal regions, precuneus, and parts of the DMN along medial PFC were the most connected nodes in the CPM model as in other tau models.

**Figure 4:**
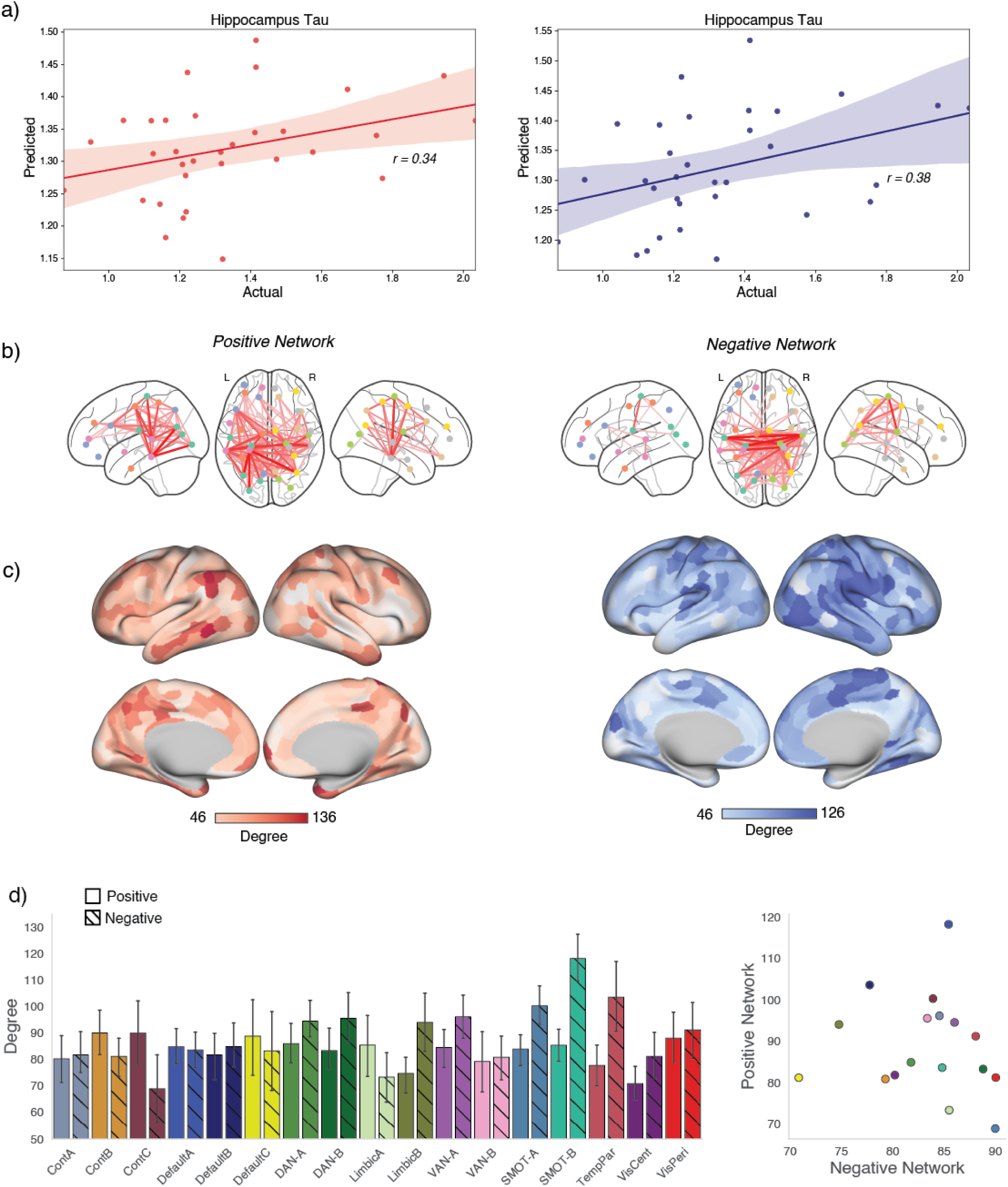
Connectome-based Predictive Modeling for hippocampal tau: a) We trained and predicted the CPM model between functional connectivity and the tau concentrations in the hippocampus within the carriers in the COLBOS dataset. b, c, d) We saw similar ROI contributions to model predictions as seen for earlier models for both positive and negative network models.

**Figure 5:**
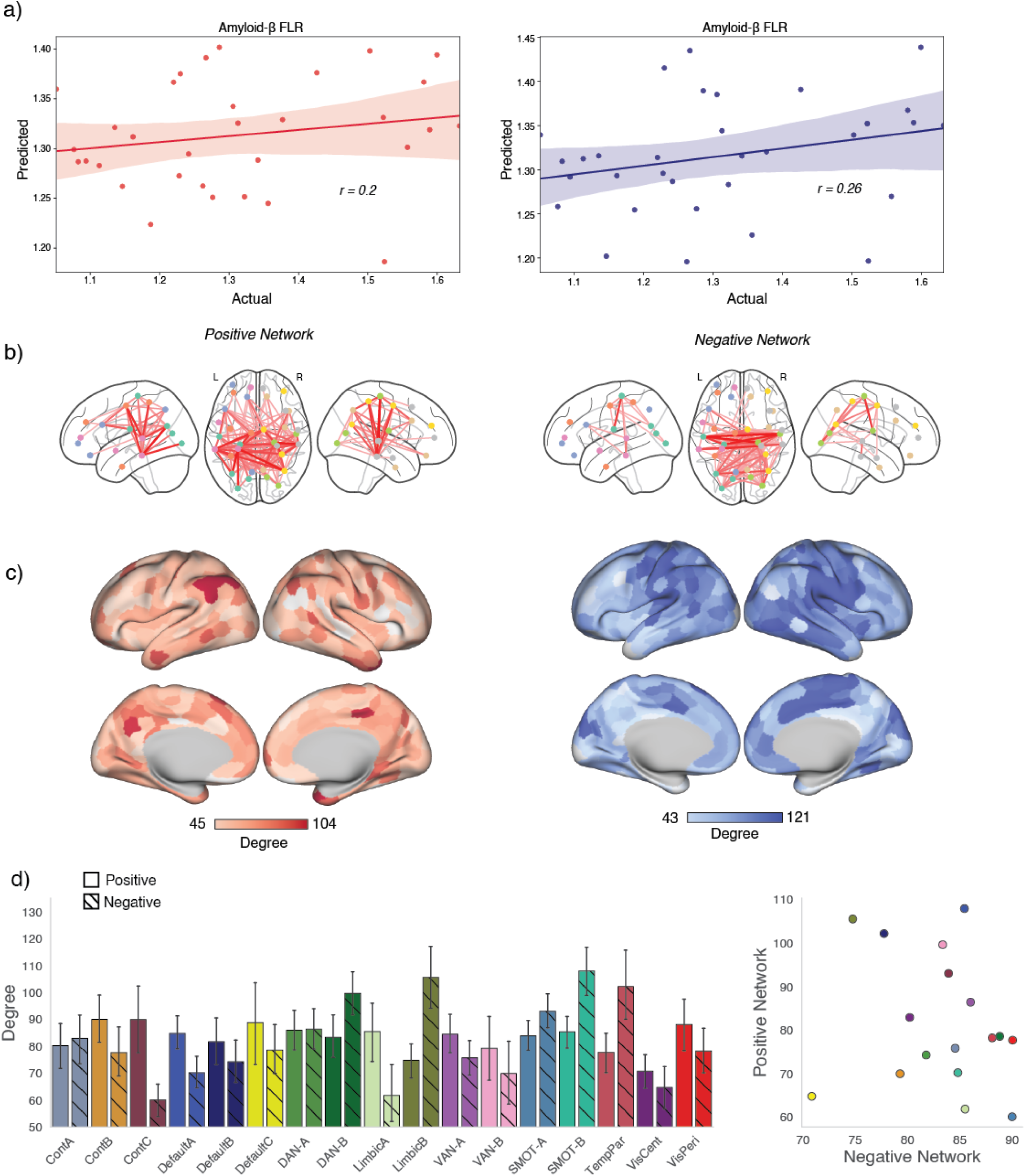
Connectome-based Predictive Modeling for amyloid-β: a) We trained and predicted the CPM model between functional connectivity and the amyloid-β concentrations in the FLR cortex within the carriers in the COLBOS dataset. b, c, d) Again, as seen for tau CPM models, connectivity between IPL, DAN, IT was the most predictive for positive networks and somatomotor, pre-SMA, precuneus, temporal cortex connectivity was driving predictions for negative networks.

**Figure 6:**
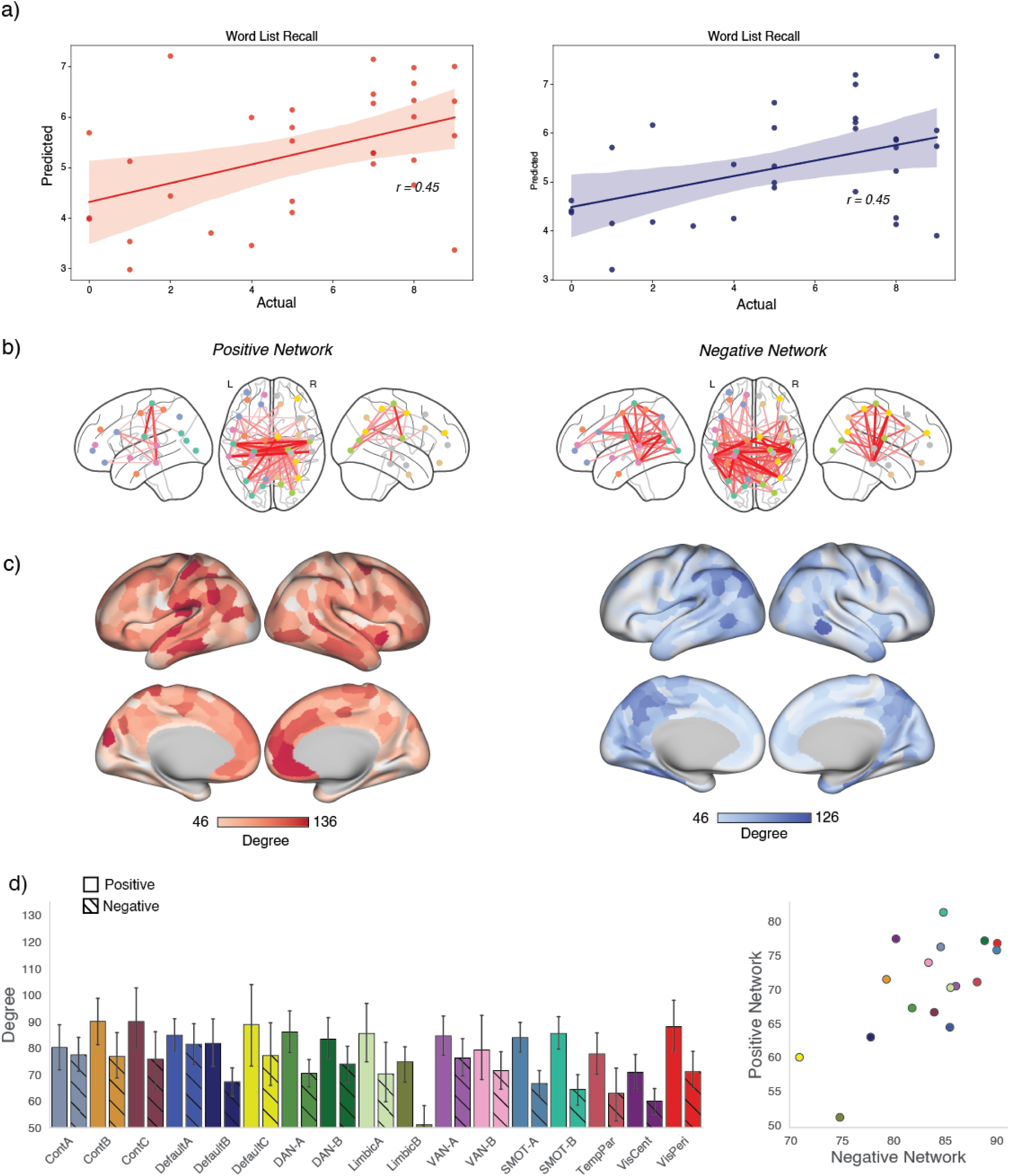
Connectome-based Predictive Modeling for cognitive scores: a) We trained and predicted the CPM model between functional connectivity and word list recall total behavioral score within the carriers in the COLBOS dataset. b) We see switched trends for the positive and negative networks for the word list recall values as subjects with higher tau/amyloid-β concentrations had lower cognitive scores. c, d) We saw the contributions of somatomotor connectivity along with temporal cortical regions and mPFC for the positive networks and IPL, IT and posterior cingulate cortex connectivity contribute towards the prediction using negative network models.

**Figure 7:**
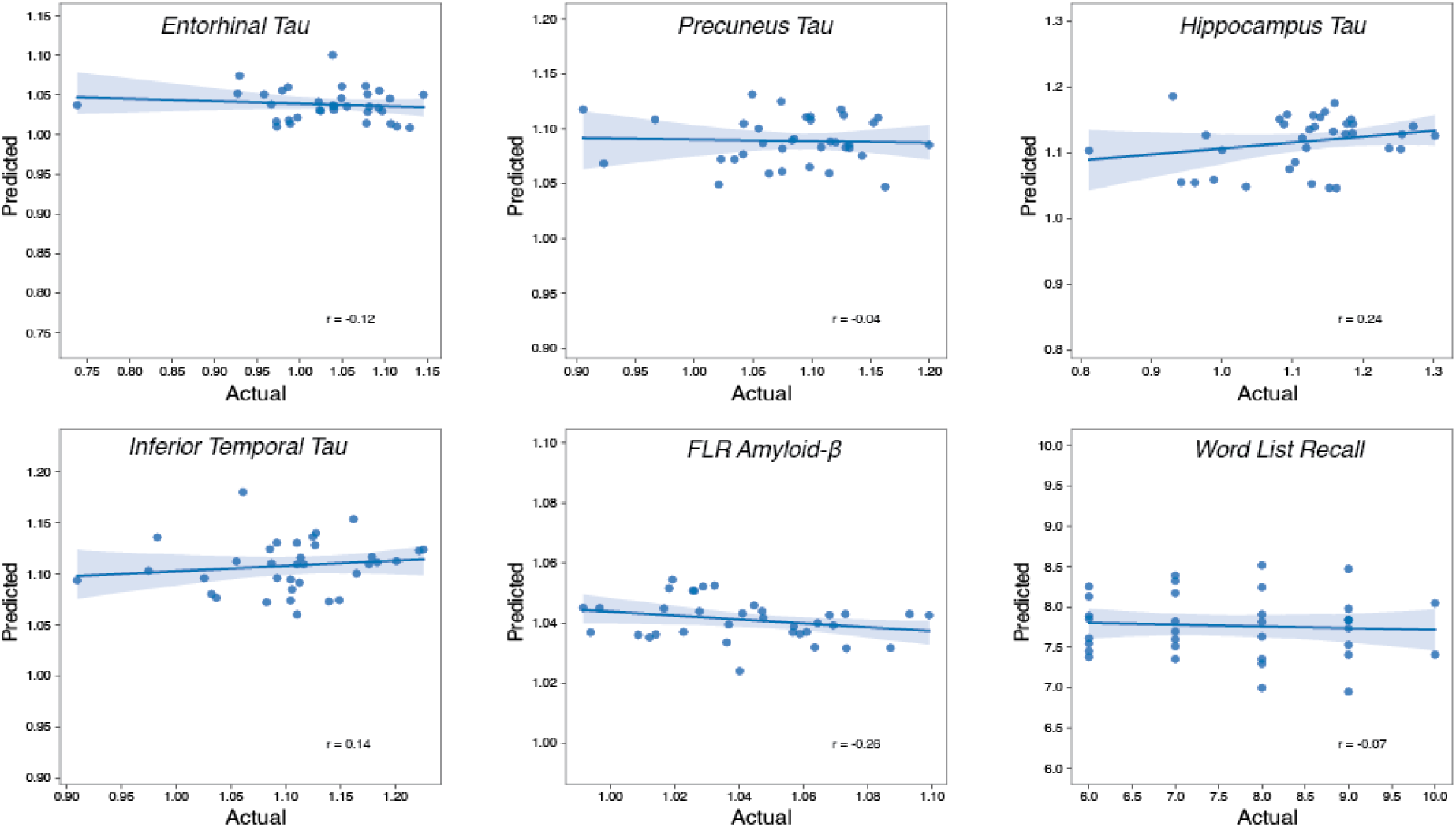
Connectome-based Predictive Modeling in *PSEN1* non-carriers: We applied the CPM modeling between functional connectivity across parcels and the tau concentrations in the entorhinal, precuneus, hippocampus and IT. The amyloid-β in FLR cortex and word list recall total cognitive scores in the non-carriers within the COLBOS datasets were also investigated. We find that the model is weakly predictive for hippocampal values but not any others.

**Figure 8:**
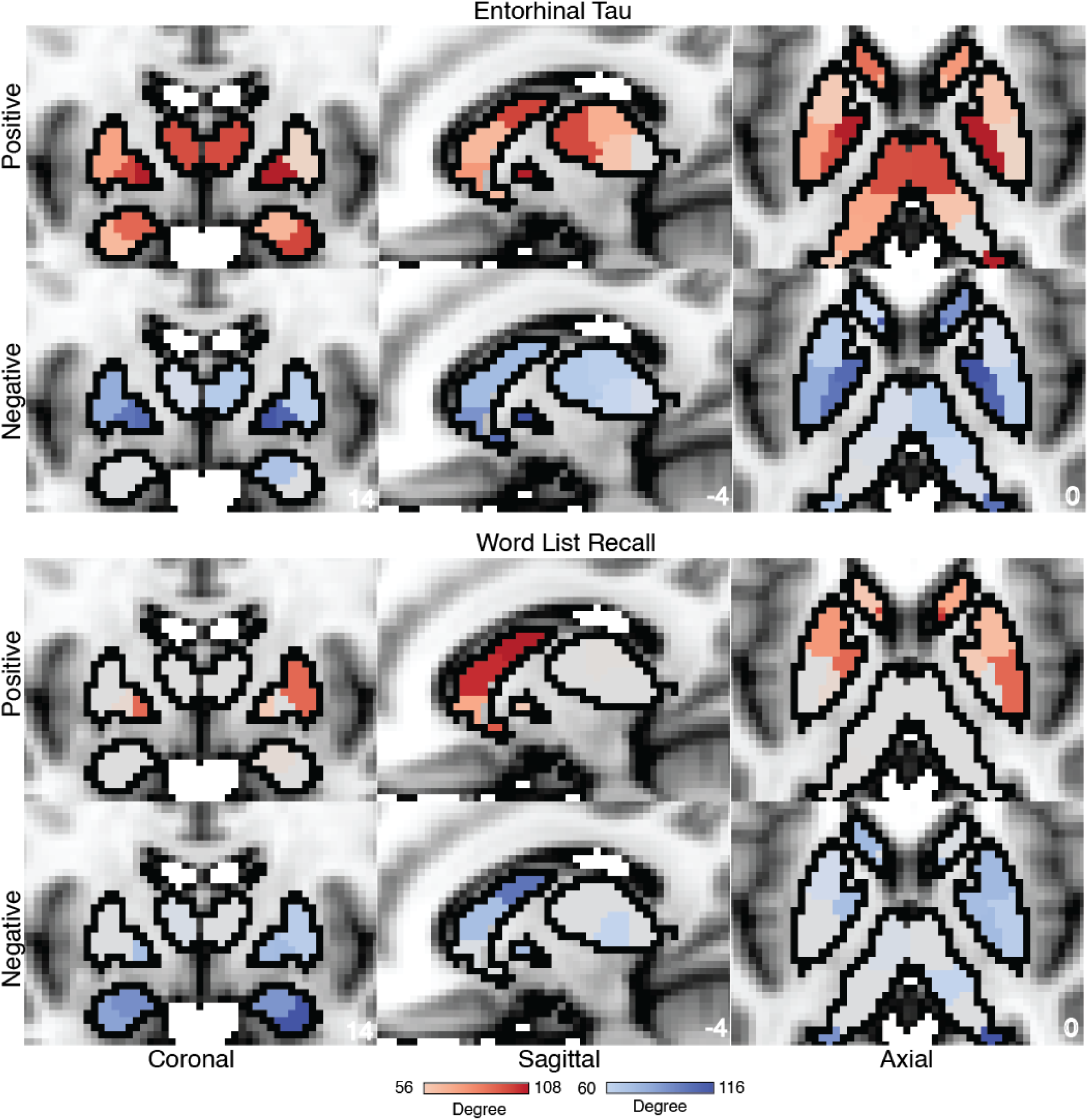
Subcortical contributions to the predictive model: Degree centrality of the predictive model (separately for positive and negative connections) for entorhinal tau and word list recall values trained for *PSEN1* carriers. We see a pronounced contribution of anterior thalamus, globus pallidus, and dorsal striatum for the model predictions for entorhinal tau values. For word list recall values, we find a stronger role for dorsal striatum and parts of amygdala as the most significant contributors to models’ predictions.

In our CPM models trained for various tau and amyloid-β measures across positively correlated networks, the most predictive ROIs included inferior parietal lobule (IPL) region within the DAN, DMN, cognitive control networks, and medial prefrontal cortex (mPFC) along with temporal pole. The somatomotor network, supplementary motor area (SMA) and dorsomedial prefrontal cortex (dPFC) were the strongest predictors in negatively correlated networks. We also analyzed the subcortical seeds that contributed most to the model’s success and identified the anterior thalamus, globus pallidus, and dorsal striatum. These findings suggest that the disruption of brain connectivity impacts networks across the cortical hierarchy from somatomotor regions to association cortical regions along with distributed brain networks. These findings align with the existing literature describing how tau and amyloid-β accumulate in the brain [4,5].

## 4. Discussion

MRI modalities allow for a rapid non-invasive test for brain health and function, but the utility of fMRI for translational purposes has been limited. In this study, we sought to establish the relationship between functional connectivity and the accumulation of tau tangles and amyloid-β plaques in carriers of an ADAD mutation. We utilized Connectome-based Predictive Modeling to successfully predict tau and amyloid-β concentrations in *PSEN1* E280A carriers. The connectivity between the regions in the IT and the IPL in the DMN, DAN and somatomotor network were the most predictive aspects of the networks. This suggests that these edges are the most influenced by the change in tau values and can be useful as potential biomarkers for AD diagnosis in the future. Further node-level degree analysis of these models showed strong predictability of DMN nodes in IPL, mPFC, dPFC, and somatomotor regions in the model predictions suggesting that large-scale networks are disrupted due to tau and amyloid-β spread across the brain. Prior studies have shown that greater tau accumulation decreases the within-network connectivity DMN regions like precuneus and medial PFC but increases the within and across-network connectivity in anterior cingulate and retrosplenial cortex [9]. Our findings complement this study, and we now highlight which functional connections were the most predictive of tau and amyloid-β burden not limiting to the regions with the most changes in within and across network connectivity. We also identified that subcortical ROIs like the dorsal striatum, anterior thalamus and globus pallidus are strong predictors of tau deposition. This could be due to disruption in large-scale association networks which are distributed across parts of the basal ganglia and thalamus [43,44]. Our models also significantly predicted behavioral scores like the word list recall, total corrects and MMSE scores. The network maps between the positive and negative networks were switched due to the directionality of the scores (lower scores represented higher cognitive decline).

The study was limited to the analysis of tau tangles and amyloid-β plaque deposits in the brain and its prediction using resting-state functional connectivity. We did not analyze the change across longitudinal sessions, which we can expand on in future studies. The resting-state data was collected across two runs of around six minutes each. Though the data is usually sufficient, studies have shown that for predictive purposes, more than twenty minutes of data give more stable network estimations and hence better results [11,16,45]. Another limitation of this study is the use of global parcellations to define the nodes used to create brain networks. Although these parcellations are accurate to a large degree, recent advances in precision imaging have shown that the inter-subject variability might contribute to the loss of important individual features critical to the prediction of the tau and amyloid-β pathology. Although we did not account for sex in the model, earlier studies have shown differences in clinical symptomatology and tau/amyloid-β pathology across sex with boys showing faster decline and poor cognitive abilities as compared to girls [46]. Future models with larger samples should consider this.

While CSF- and blood-based biomarkers demonstrated successful disease staging in AD [47,48], functional MRI-based approaches provide an *in-vivo* localization of AD-related pathological processes and was successfully used to anticipate tauopathy progression [49–51], making both approaches complementary.

The present CPM analysis utilizes an OLS approach. Future analyses may benefit from a regularized regression approach or from non-linear methods to get a better fit between the connectivity and tau/amyloid pathology. Still, CPM provides a useful model for prediction within the COLBOS dataset and could be applied to other ADAD studies. It helps us to understand network connectivity change related to AD and may help in early diagnosis. Studies have also shown that AD progression is heterogeneous across subjects, with at least four different kinds of trajectories [25,27,49–52]. Further research is required to train models that can predict these different progressions across tau and amyloid deposition. We also did not analyze the impact of neuroinflammation of functional connectivity which some recent studies pointed out [53].

Another potential limitation of the present network approach is that connectivity estimates can vary substantially with preprocessing strategies. Research has shown that physiological noise can drive resting-state functional connectivity [54]. Global signal regression can also “drive” certain networks, thus, giving wrong network estimates [55]. Multiple strategies have been suggested to deal with overall signal noise in the resting-state fMRI data [56–58]. Freely available tools and pipelines have standardized preprocessing approaches [31,59]. However, ,the effect of different variables of the preprocessing pipeline, like distortion and motion correction, or and global signal regression on network estimates, is still lacking and requires further research. Specifically, how the preprocessing strategies for populations with AD can impact connectivity estimates and model predictability would require further investigation. Though scientists have pointed out that using such Brain Wide Association Studies (BWAS) for small populations could affect results and reproducibility [23], proper study design allows reproducible and robust predictions [24]. Here, we analyzed brain-brain correlations which we think are more robust, and described important aspects of the brain’s organization principles [18].

The ADAD population typically develops clinical symptoms at an earlier age, displaying phenotypic changes around the age of 35-40 with dementia setting around mid-40s. The near certainty of AD development offers a unique opportunity to examine the progression of AD over many years prior to the emergence of clinical symptoms. While ADAD patients potentially provide a window into the development of AD in other patient groups, notable differences between ADAD and other forms of AD exist. Our study also found that connectivity-based models better predicted tau burden than amyloid-β deposits. This might be due to the rapid changes in tau accumulations in the ADAD cohort resulting in stronger connectivity disruptions. And tau accumulations may be more associated with connectivity changes than with amyloid-β deposits. Tau increase in *PSEN1* carriers is stronger than in late-onset AD patients in line with the rapid decline experienced in the subjects [5]. Studies also pointed towards differences in tau and amyloid progression across the various forms of AD [4,27,60]. There might be an exponential tau accumulation in *PSEN1* carriers which would suggest cascading effects across brain regions which requires further investigation. Future models should incorporate such nonlinear changes in tau accumulation and associated functional connectivity alterations.

Possible future studies of interest include models that can predict growth and changes in tau fibrillation and amyloid-β plaques based on current resting-state connectivity measures. We can also utilize an individualized parcellation-based method to improve the estimations of subject-level brain connectivity that would better capture the state of tau/amyloid-β pathology.

In summary, this study finds strong correlation between resting-state functional connectivity measures and both tau/amyloid-β concentrations and behavioral scores in a unique dataset of subjects with ADAD. Such connectivity models provide a complementary approach to PET scans could and could be used in the clinical setting for an early diagnosis of and monitoring of AD.

## Data Availability

The data in the study are available on request from the corresponding author Y.T.Q.

## Acknowledgements

The authors thank the Colombian Families for their commitment and dedication to the COLBOS project.

